# Automated microfluidic electrochemical biosensor for the detection of immune-mediated thrombotic disorders

**DOI:** 10.1101/2025.05.15.25327633

**Authors:** Diana F. Cedillo-Alcantar, Adam Kanack, Seonhwa Lee, Alan M. Gonzalez-Suarez, Kihak Gwon, Emily Mauch, Thi Thanh-Qui Nguyen, Anand Padmanabhan, Alexander Revzin

## Abstract

Vaccine-induced immune thrombotic thrombocytopenia (VITT) is a new disorder that emerged in the wake of COVID-19 vaccination. It is a rare but life-threating condition that requires aggressive course of treatment to improve patient outcomes. To date, there has not been an effective diagnostic assay for detecting VITT. Instead, definitive diagnosis requires satisfying several criteria including history of recent vaccination, platelet count, positive ELISA result for a closely related thrombotic disorder, heparin-induced thrombocytopenia (HIT) and PF4-dependent functional assays. Our study describes a technically simple antigenic assay for direct diagnosis of autoimmune antibodies (Abs) associated with VITT. We first show that cross-linked platelet factor 4 (PF4) represents an antigenic target specific for VITT Abs. We then incorporate this antigenic target into a microfluidic electrochemical biosensor and demonstrate specific and sensitive detection of VITT Abs in a fully automated manner while using microliter volumes of patient sera. We tested 51 patient samples using the microfluidic electrochemical biosensor and demonstrated 100% sensitivity and specificity for VITT sera compared to healthy controls and HIT patients.

## INTRODUCTION

Vaccine-induced immune thrombotic thrombocytopenia (VITT) is a rare but severe thrombotic disorder most frequently caused by an immunologic reaction to adenoviral vector-based COVID-19 vaccines (*1, 2*), and rarely after other types of non-COVID-19 vaccinations (e.g., an HPV vaccine). VITT is characterized by the production of pathogenic antibodies (Abs) with specificity for platelet factor 4 (PF4) (*3*) that trigger platelet activation and cause patients to experience thrombocytopenia and thrombosis (*2, 4*). These features closely resemble other thrombotic disorders mediated by anti-PF4 Abs, such as heparin-induced thrombocytopenia (HIT), a disorder that can occur in patients treated with heparin(*5*). In some cases, individuals can develop thrombosis and/or thrombocytopenia spontaneously (without proximate heparin exposure) due to antibodies that look like VITT antibodies; this condition is referred to as “Spontaneous VITT”(*6, 7*) as well as the more recently described monoclonal gammopathy of thrombotic significance (MGTS)(*8–10*). MGTS antibodies targets can be complex with generation of antibodies that are HIT-like (*13*), VITT-like (*14*), or neither HIT or VITT-like (*15*).

HIT and VITT are associated with mortality rates of about 10% (HIT) and about 20% (VITT) without early detection and prompt management. It can be challenging to differentiate HIT antibodies from VITT and spontaneous VITT antibodies. The anti-PF4 antibodies that cause VITT are not well characterized, but they can be detected with at least two types of laboratory assays: antigen-based immunoassays, in select cases, platelet activation-based functional testing, which is technically challenging and performed in specialized reference laboratories(*16*). Enzyme linked immunosorbent assays (ELISAs) employing PF4/polyanion complexes as an antigenic target are typically used as a frontline test for detecting anti-PF4 Abs associated with HIT, However, these antigen-based assays are non-specific, detecting antibodies associated with VITT and HIT (*16*), as well as non-activating non-pathogenic PF4-polyanion ELISA-positive antibodies that are present in more than 4% of the general population with no obvious pathology (*17*), and non-activating, non-pathogenic anti-PF4 antibodies that are commonly found in heparin-exposed individuals(*18*). Rapid/automated HIT immunoassays have extremely poor sensitivity for detection of VITT antibodies (*19*). None of the currently approved assays possess specificity for VITT Abs (*20, 21*). Various platelet function assays have been developed to assess aggregation, agglutination or degranulation (e.g. serotonin release) (*22–27*). Only PF4-enhanced washed platelet assays reliably recognize VITT Abs but are less suited for the frontline setting, given extremely limited availability due to requirement for fresh donor platelets and complex techniques, and are generally recommended only for confirming pathogenicity of anti-PF4 VITT Abs following a positive ELISA test (*28*). Therefore, technically-simple, yet effective diagnostic assays capable of distinguishing VITT Abs from closely related HIT Abs are lacking and are sorely needed given the more aggressive nature of VITT and the criticality for timely treatment.

The objective of our study was to address this need by developing an immunoassay for specifically yet sensitively detecting VITT Abs. We wanted this immunoassay to be rapid and automated to make it suitable for a frontline or point-of-care setting, given that VITT frequently presents in the out-of-hospital environment in otherwise healthy individuals who get vaccinated. Our team has recently described an electrochemical immunoassay using gold nanoparticles (AuNP) functionalized with Abs and doped with metal ions to detect surface markers on extracellular vesicles (*29, 30*). These nanoparticles (also called immunoprobes) offered two key advantages enabling sensitive detection of a target analyte: 1) each AuNP is decorated with multiple Abs, creating multivalent interactions that enhance target affinity, and 2) each AuNP is estimated to contain 10^4^ metal ions, amplifying electrochemical redox signal (*30*). We reasoned that automating the workflow of the electrochemical immunoassay while using minute quantities of sample will make the assay particularly well-suited for the point-of-care setting. Microfluidic devices with computer-controlled microvalves offer exciting possibilities for pumping and routing microliter volumes(*31–34*). Our team has employed such devices previously to isolate plasma and detect biomarkers based on 5 µL of whole blood(*35*) and to perform high-throughput serology assays (*36*).

In this study, we sought to combine an electrochemical immunoassay with microfluidic automation to develop a rapid, sensitive, and specific assay for detecting VITT Abs. The specificity of the assay was enabled by a novel observation that cross-linked PF4 (c-PF4) molecules represent antigenic target specific for VITT Abs while uncross-linked PF4 molecules complexed with heparin are reactive to either VITT or HIT Abs. This observation was leveraged to design a sensing strategy shown in Figure 1A, where Au electrodes were functionalized with heparin/PF4 complexes, incubated with patient sera and then labeled with immunoprobes. Binding of VITT or HIT Abs to antigenic target was determined electrochemically.

**Fig. 1.**
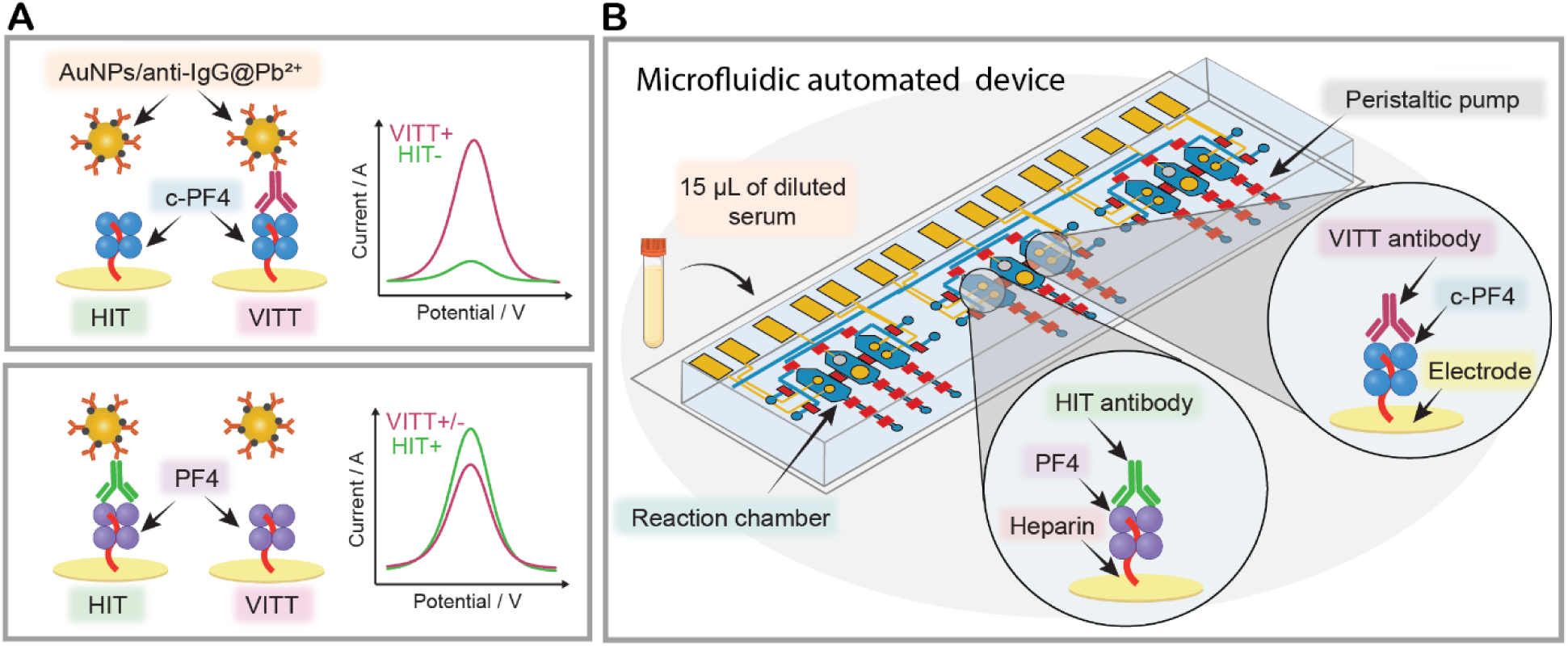
Overview of the microfluidic electrochemical biosensor. (**A**) A sensing strategy developed in this study involves immobilizing thiolated heparin /PF4 complexes on Au electrodes. Some of the electrodes are functionalized with cross-linked PF4 (c-PF4) molecules that are specific to VITT Abs while other electrodes carry uncross-linked PF4 molecules reactive with either VITT or HIT Abs. Using these antigenic targets in the same diagnostic platform allows us to distinguish the presence of VITT or HIT Abs in a given serum sample. Labeling electrodes with gold nanoparticles carrying anti-human IgG and Pb^2^ (AuNPs/anti-human IgG@Pb^2^) is followed by voltammetry to produce electrochemical signals proportional to the concentration of surface bound anti-PF4 Abs. **(B)** Electrodes carrying biorecognition molecules are incorporated into a microfluidic device. The device (microfluidic electrochemical biosensor) contained multiple microchambers to enable parallel analysis of patient sera as well as positive and negative controls. Microvalve integration in the device allowed full automation of labeling-incubation-washing cycle while using only 15 µL of diluted patient sera.

Importantly, sensing electrodes were incorporated into a series of valve-controlled microfluidic chambers and were used to determine VITT vs. HIT status based on an input of a small volume of serum, 15 µL, in a fully automated fashion (see Figure 1B).

## RESULTS

### Modifying PF4 to tune antigenic specificity to VITT Abs

A key goal of this study was to develop antigenic targets to VITT Abs to be able to differentiate them from closely related HIT Abs. PF4 is the antigenic target for both Ab types (see Figure S1A for amino acid sequence). However, there are indications that each Ab type recognizes different epitopes (*37*). It has been established that HIT Abs recognize PF4 complexed to heparin or other polyanions (*38*). As expected, a commercial ELISA based on PF4/polyvinylsulfonate complexes did not give us the ability to discriminate between HIT and VITT (Figure 2A). Our team has previously shown that VITT Abs interact with uncomplexed PF4 and that uncomplexed PF4 may be used to construct ELISA for VITT detection (*3*). However, this past study was carried with a cohort of only 6 VITT patient. When testing a larger cohort of samples (11 VITT and 21 HIT) using ELISA with uncomplexed PF4 as antigenic target we did not observe significant difference between the two groups (Figure 2B). We then explored an approach of cross-linking PF4 to alter its conformation and make it more specific to VITT Abs. Pursuit of this approach was catalyzed by our observation that gentle cross-linking of PF4 molecules with 1-ethyl-3-(3-dimethylaminopropyl) carbodiimide (EDC) provided sufficient epitopes for VITT Ab binding (see Figure S1A for description of cross-linking). In fact, optimal cross-linking conditions did not alter PF4 oligomerization. As shown by SDS-PAGE and ultracentrifugation (see Figure S1(B,C)), cross-linking of PF4 yielded a mixture of oligomeric species, with ∼81.2% of PF4 present as tetramers.

**Figure 2.**
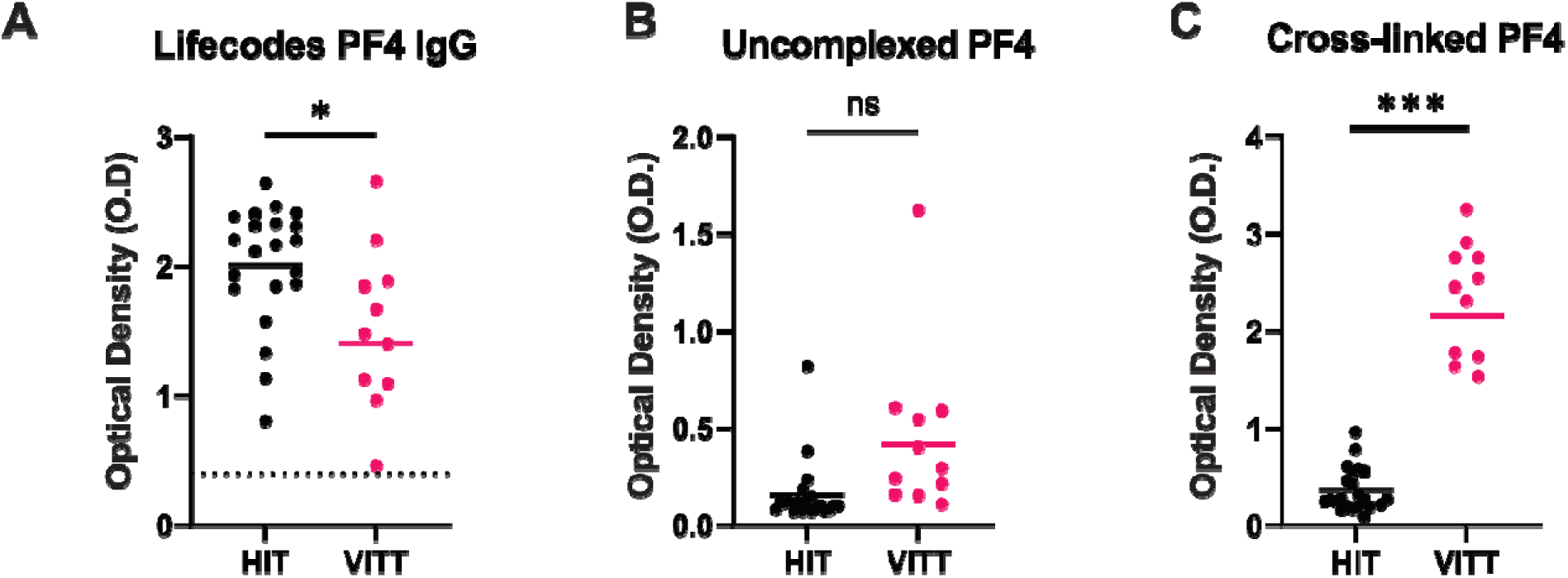
Designing antigenic target to improve specificity for VITT Abs. A cohort of patients with clinically confirmed HIT (n=21) or VITT (n=11) were tested with ELISAs utilizing (**A**) PF4/polyanion, (**B**) uncomplexed PF4 or (**C**) cross-linked PF4 targets, to compare the sensitivity and specificity of each antigenic target for detecting HIT or VITT anti-PF4 antibodies among a cohort of patient samples.

Importantly, when cross-linked (c)-PF4 was used as antigenic target in an ELISA assay, VITT sera samples produced significantly higher signals compared to HIT sera (see Figure 2C). This supported our hypothesis that cross-linking may change conformation of PF4 and improve specificity for VITT Abs. The work is ongoing to improve understanding of how the process of cross-linking exposes epitopes and improves VITT Abs binding. In the meantime, we wanted to leverage this exciting observation toward constructing a technically-simple VITT diagnostic assay suitable for deployment in the frontline or point of care setting.

### Design of the electrode interface to enable sensitive and specific detection of HIT and VITT

Building on the observation that cross-linking of PF4 improves immunoassay sensitivity toward VITT Abs, we proceeded with development of electrochemical immunoassays. Given that heparin is commonly used in a complex with PF4 for detecting anti-PF4 Abs, we wanted to explore a strategy for immobilizing heparin/PF4 complexes on an electrode surface. To achieve this, heparin was functionalized to contain thiol groups because thiolated molecules form strong, stable bonds with Au substrates. Subsequently, surface plasmon resonance (SPR) was used to characterize and optimize the assembly of the biorecognition layer comprised of heparin-thiol and either PF4 or c-PF4. The use of SPR allowed us to optimize construction of the biorecognition layer on the Au substrate and apply the optimized protocol for electrochemical microfluidic biosensing experiments described later in the paper. Figure 3A shows a representative SPR sensogram depicting heparin-thiol binding and poly(ethylene glycol) (PEG)-thiol blocking steps. The solution concentration of heparin-thiol leading to the maximal capture of PF4 molecules was optimized (Figure S2) and was found to be 0.25mg/ml. After creating the Hep-SH/PEG-SH layer, the SPR chips were exposed to either PF4 or c-PF4 to create a complex for recognizing HIT or VITT Abs. These surfaces were then incubated with HIT or VITT patient serum. As a final step, anti-human IgG Abs were introduced into the SPR flow cells to confirm the presence of human IgG Abs on the surface. Given that serum contains multiple proteins capable of interacting with heparin (*39*), labeling with anti-human IgG Abs was used to reveal the presence of IgG Abs rather than non-specific serum protein interactions. SPR results confirmed the successful construction of a biorecognition layer comprised of Hep-SH/PF4 or Hep-SH/C-PF4 complexes and the capture of Abs from HIT or VITT serum followed by the binding of anti-human IgG to the HIT and VITT Abs (Figure 3A-B).

**Figure 3.**
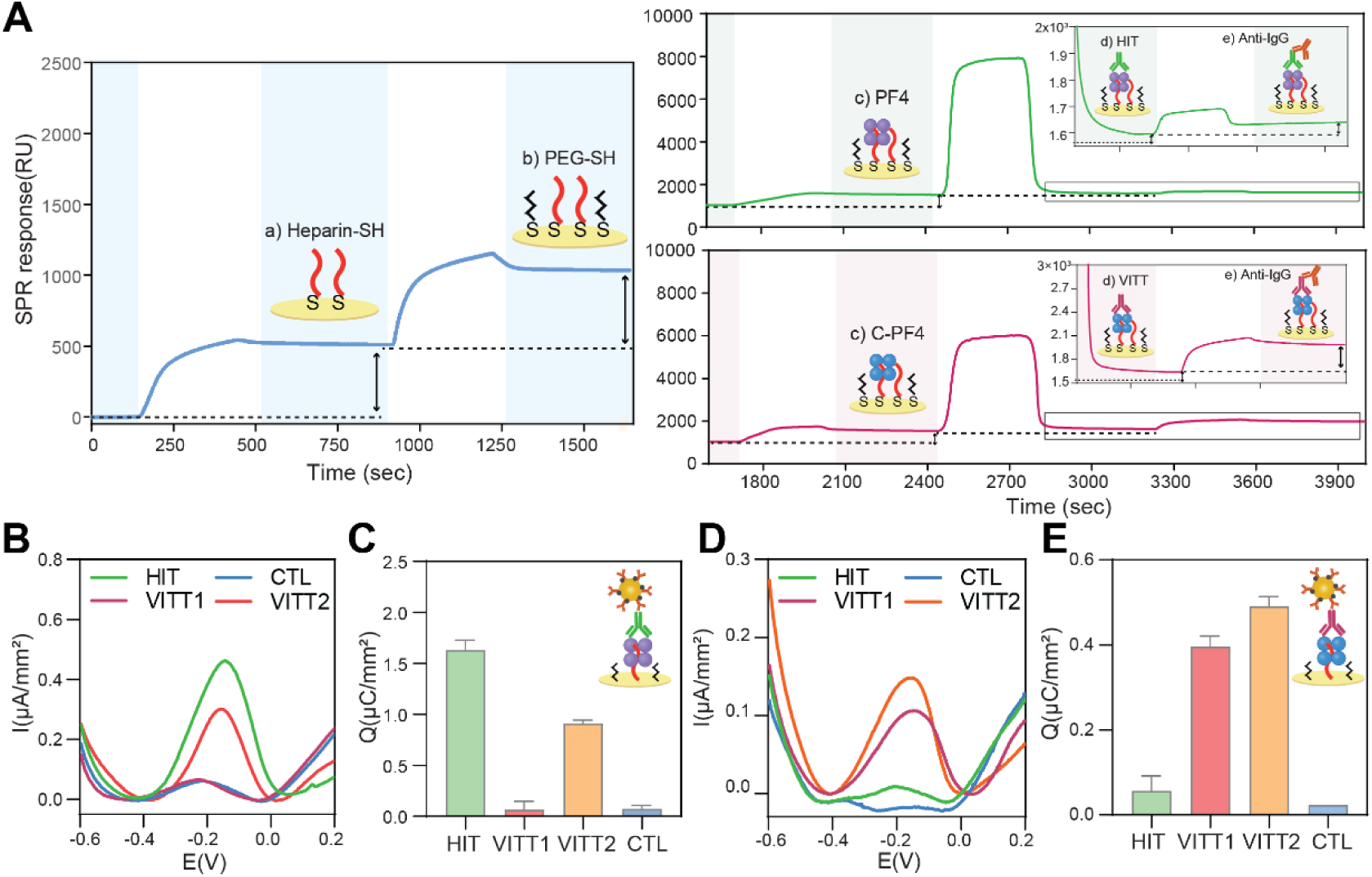
Assembly of the biorecognition layer and analysis of biosensor specificity toward HIT or VITT Abs. **(A)** SPR analysis of steps involved in the assembly of the biorecognition layer: a) Immobilization of heparin-SH, b) passivation with PEG-SH and c) binding of either PF4 or c-PF. PF4-containing biorecognition layer was challenged with HIT serum while c-PF4 layer was exposed to VITT serum. Anti-human IgG Abs were used enhance the signal. **(B)** SWV signals from electrodes functionalized with PF4 and exposed to either healthy serum (blue), HIT serum (green), VITT1(red) or VITT2 (orange) serum. This result illustrates that HIT sera produced higher signals than control but that some VITT samples (e.g. VITT2) produced high signals as well. Thus, the ability to discriminate HIT from VITT was limited when using PF4 as antigenic target. (**C**) Normalized charge values for experiment described in part B. (**D**) SWV signals for electrodes functionalized with c-PF4 and challenged with healthy (blue), HIT (green), VITT1 (red), or VITT2 (orange) patient sera. This result demonstrated specificity for VITT Abs. (**E**) Normalized charge values for experiment described in part D.

Having characterized the construction of the biorecognition layer and binding of HIT or VITT Abs to this layer, we designed the electrochemical biosensor integrated into a simplified microfluidic device. As shown in Figure S3, this device was fabricated in poly (dimethyl siloxane) (PDMS). It contained six independent microfluidic compartments, each with a microchamber of 1 µL and two Au working electrodes (600 µm diameter) patterned on glass. The electrodes were functionalized using a sequence of steps outlined in Figure 3A. However, unlike SPR analysis where anti-human IgG Ab molecules were used, electrochemical detection was carried out with immunoprobes (AuNPs/anti-human IgG@Pb² ). As shown in Figure S3A, square wave voltammetry (SWV) analysis initially revealed a high background signal in the control sample. The total charge (Q) was calculated from the SWV curves, and the signal-to-noise ratio (Q_HIT_/Q_CTL_) was found to be 2.6 (Figure S4B). To determine whether the signal-to-noise ratio could be improved through better passivation of the electrodes, we evaluated experimental parameters that included three different passivation agents and various incubation times. These experiments revealed that the suboptimal S/N was attributable to the non-specific interactions between the electrode and the immunoprobes (Figure S5, S6). Passivating the electrodes with PEG-SH for 1h minimized non-specific binding of immunoprobes as confirmed by SPR (see Figure S6, S7) and allowed us to increase S/N of electrochemical detection to 8.8 for the same serum sample that previously yielded an S/N of 2.8 (Figure S3). Next, we evaluated the specificity of the electrodes functionalized with PF4 and c-PF4. Electrodes containing either target antigen were challenged with control, HIT, or VITT serum. Figure 3B-E summarizes the electrochemical responses for the different antigen/serum combinations. These results confirm that our electrochemical biosensor was specific to either HIT or VITT Abs depending on the antigen used.

### Design and operation of the automated microfluidic electrochemical biosensor

Given the ultimate goal of eliminating manual handling, we proceeded to automate the assay workflow in the microfluidic device. The design criteria for this device were 1) to enable the detection of both HIT and VITT, 2) to incorporate positive and negative controls for signal normalization, and 3) to utilize a minimal volume of serum for analysis. Figure 4A-B shows the device developed to accomplish these goals. The device contained three analytical units, each comprising two sensing compartments and one compartment housing reference (RE) and counter electrodes (CE). The device also contained a series of pneumatic microvalves used for liquid routing and pumping. Figure 4B offers a close-up view of one sensing unit. As may be noted, each sensing compartment contained two working electrodes. Figure 4C illustrates the configuration of the three analytical units, where each sensing compartment contains either PF4 or c-PF4. The first unit was designed for serum sample analysis, the second for positive controls specific to HIT and VITT, and the third for negative controls (CTL). The sensing and RE/CE compartments were connected by pneumatically controlled microvalves that were kept closed to isolate working electrodes during serum incubation and immunoprobe labeling steps and were opened only when making electrochemical measurements. This allowed us to keep the RE/CE compartment pristine and unaffected by serum before electrochemical measurements.

**Fig. 4.**
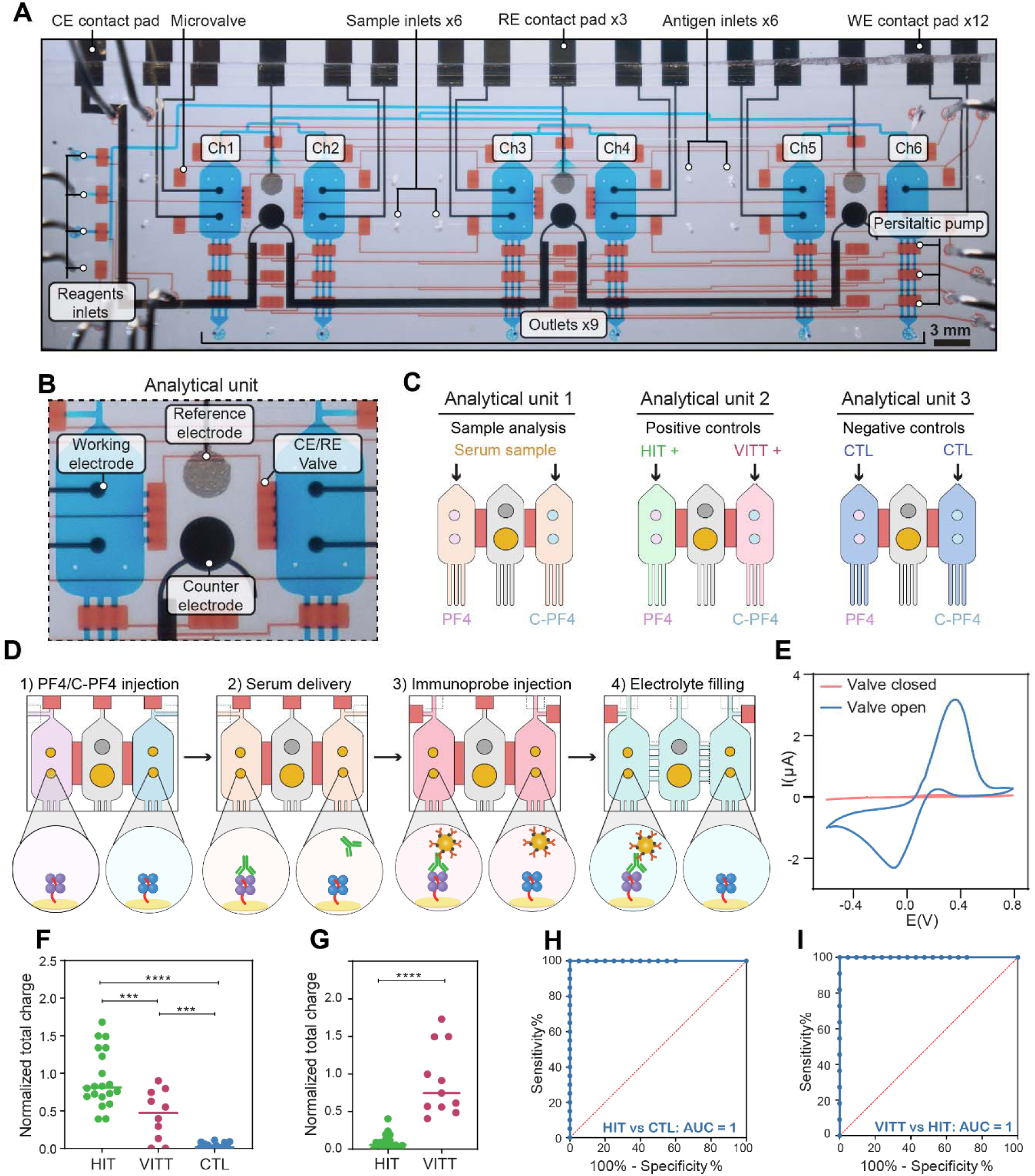
Automated operation of the microfluidic device and analysis of patient sera. (**A**) Photograph highlighting device components where the valve (control) layer filled was filled with blue dye and the flow layer was filled with red dye. (**B**) Close-up view of one analytical unit comprised of three microchambers. Two microchambers contained working electrodes and were flanking a chamber with reference and counter electrodes. Fluidic connection between the chambers was controlled by microvalves. (**C**) Description of how analytical units were used in an assay. (**D**) Illustration showing steps of the assay: 1) immobilization of antigen targets, 2) introduction of serum samples, 3) labeling with immunoprobes and 4) actuation of microvalves to connect working electrodes to counter and reference electrodes for electrochemical measurements. (**E**) [Fe(CN)6]^4-/3-^ cyclic voltammetry demonstrating that the connection to counter and reference electrodes was established only with opened microvalves. (**F**) Analysis of patient sera samples in a microfluidic electrochemical biosensor with PF4 as antigenic target. HIT samples produced significantly higher signals than VITT samples and controls. (**G**) Analysis of patient sera in a microfluidic biosensor with c-PF4 as antigenic target. VITT samples produced signals that were significantly higher compared to HIT sera. (**H, I**) ROC curves demonstrating 100% sensitivity and specificity for detecting HIT and VITT evaluate.

The automated assay workflow is shown in Figure 4D and Movie 1. It starts by isolating the CE/RE compartments from sensing compartments. The biorecognition layer was constructed on the working electrodes using either PF4 or c-PF4. Next, a test serum sample as well as positive and negative controls are pumped into the sensing compartments for 10 min. On-board peristaltic pumps allow us to use as little as 15 µL of patient sample during testing. After washing with PBS-Tween (PBS-T), immunoprobes were flowed into all the chambers for 10 min and then washed again with PBS-T and DI water. Subsequently, an electrolyte (acetic acid/sodium acetate buffer) is introduced into all compartments, and the valves isolating the sensing from CE/RE compartments were opened. Figure 4E describes [Fe(CN) ] ^-^/³^-^ cyclic voltammetry performed in conjunction with valve actuation and shows that the electrochemical signal was only observed when channels connecting working electrodes to CE/RE electrodes were opened.

### Performance of the microfluidic automated electrochemical biosensor

Before testing serum samples, we carried out extensive characterization of the device. First, we evaluated the electrochemical performance of each of the 12 working electrodes in the device to confirm that the location of the electrodes did not affect their properties. Figure S8(A-B) shows representative cyclic voltammetry and SWV responses of electrodes in 10 mM [Fe(CN)6]^4-/3-^ in 0.1 M KCl. As may be appreciated from these data, anodic and cathodic peak currents were similar between the working electrodes regardless of location, with a coefficient of variation (CV) of less than 5%.

We next evaluated biosensor responses to HIT-like Abs first identified in two patients diagnosed with monoclonal gammopathy of thrombotic significance (MGTS) whose monoclonal anti-PF4 Abs were de novo sequenced and produced recombinantly. These Abs provided us with a positive control for HIT detection and were used to establish analytical performance (limit of detection, dynamic range) of our electrochemical biosensor. Challenging electrodes with varying concentrations of HIT-like Abs allowed us to establish a limit of detection (0.12 µg/mL) and dynamic range (1.3 µg/mL) of the electrochemical biosensor (see Figure S9). Given that the sequence of VITT Abs has not been identified to date, similar characterization of our VITT biosensor was not possible.

### Validation of microfluidic automated electrochemical biosensor using a cohort of VITT and HIT samples

Our microfluidic automated electrochemical biosensor was validated using a cohort of 51 serum samples. Twenty samples were from patients who tested positive for HIT Abs using commercial assays, eleven were from patients whose VITT diagnosis was confirmed by platelet function assay while the remaining twenty serum samples were from healthy controls.

Electrochemical signals were recorded from two different working electrodes, and the total charge (Q) was calculated by integrating the area under the SWV curve. The normalized charge values (Q) were calculated using the formula (Q_S_-Q_NC_)/(Q_PC_-Q_NC_), where Q_S_ represents the charge of the clinical serum sample, Q_NC_ represents the charge of the negative control (control serum), and Q_PC_ represents the charge of the positive control (HIT-like Abs or VITT positive serum sample). As shown in Figure 4F, electrodes functionalized with PF4/heparin complexes showed significantly higher signals in HIT sera compared to healthy sera, as well as a significant difference between HIT and VITT sera and between VITT and control sera. These findings indicate that PF4/heparin provides some level of differentiation between HIT and VITT, although with overlapping signals in some VITT samples. This result was unsurprising given that other immunoassays relying on PF4-polyanion complexes as antigenic targets have shown a limited ability to distinguish HIT and VITT Abs (*12*). However, specific detection of VITT sera was achieved using electrodes containing c-PF4/heparin complexes. As shown in Figure 4G, VITT sera produced significantly higher signals compared to HIT sera. These results are exciting as they demonstrate, for the first time, the ability to differentiate between HIT and VITT sera samples in the same diagnostic platform.

To determine the utility of our automated microfluidic electrochemical biosensor as a diagnostic tool, we generated receiver operating characteristic (ROC) curves. ROC is used to obtain the appropriate cutoff value to establish optimal sensitivity and specificity for a diagnostic test. ROC is also used to evaluate the predictive value of a test by quantifying the area under the curve (AUC), where values above 0.9 suggest highly accurate tests to distinguish between conditions (*40*). As shown in Figure 4H, the area under the curve of ROC was 1, indicating the correctly classified diagnosis of HIT from healthy controls. The sensitivity and specificity values in the diagnosis of HIT patients were 100% at the cutoff point of 0.2484 in normalized charge. These results highlight the diagnostic accuracy of the biosensor in distinguishing HIT from controls.

Electrodes functionalized with c-PF4/heparin complexes demonstrated superior specificity in distinguishing VITT from HIT. As shown in Figure 4I, the AUC was 1, and sensitivity and specificity were 100% at the cutoff point of 0.4820 in normalized charge, indicating high accuracy to distinguish VITT and HIT. These findings reinforce c-PF4 significantly enhances specificity, making it a more reliable alternative to PF4-enhanced platelet-activation testing for accurate VITT detection. The high specificity and sensitivity of c-PF4-functionalized electrodes highlight their potential clinical relevance for improving VITT diagnosis. In addition, our data shows that use of uncross-linked PF4 functionalized heparin electrodes is highly accurate for technically simple detection of HIT antibodies.

## DISCUSSION

VITT is a rare but potentially life-threatening condition most frequently caused by adenoviral vaccines. It is manifested by production of autoimmune Abs against a platelet protein, PF4, and is associated with severe thrombotic complications. Mechanism and clinical symptoms of VITT closely resemble that of another autoimmune thrombotic disorder, HIT. Despite the similarities, VITT and HIT are treated differently. The treatment for VITT is aggressive and typically involves a combination of direct oral anticoagulant, intravenous immunoglobulin G, and potentially therapeutic plasma exchange for refractory cases. For HIT, typically the treatment entails choosing a non-heparin alternative anticoagulant. The ability to sensitively diagnose VITT Abs and differentiate them from HIT Abs is currently lacking which complicates/delays treatment. More recent approaches to specifically detect VITT are evolving, however, they require access to major pieces of equipment (*41*).

Our study addresses the dearth of diagnostic assays for VITT, particularly ones that can be performed in the field. It describes the development of an automated microfluidic electrochemical biosensor designed for detection of HIT and VITT Abs using only 15 µL of serum. Specificity of the biosensor is ensured by a novel biorecognition layer containing either Heparin-SH/PF4 or Heparin-SH/c-PF4 complexes. These biorecognition molecules are assembled on miniature electrodes and integrated into a microfluidic device. The device also incorporates computer-controlled microvalves for routing and pumping microliter volumes in a fully automated manner. The microfluidic electrochemical biosensor included multiple microchambers to parallelize testing of serum samples as well as positive and negative controls.

Looking ahead, we envision this device being deployed at the point of care, facilitating the timely and accurate diagnosis of VITT.

## MATERIALS AND METHODS

### Study design

This study was designed to address the need for sensitive detection and differentiation of VITT and HIT antibodies. The study design was based on the following criteria: 1) the development of specific and sensitive assays using two biorecognition layers with target antigens for each thrombotic condition and 2) the integration of a nanoparticle-enabled electrochemical immunoassay into an automated microfluidic device to eliminate manual handling and reduce serum volume requirements for limited sample availability, particularly for VITT. The sample size for this study was calculated using MedCalc statistical software based on the area under the ROC curve (AUC). For differentiating VITT from HIT using cross-linked PF4, we assumed an AUC of 0.95, a type I error (alpha, significance) of 0.005, a type II error (Beta, 1-power) of 0.005, a null hypothesis value of 0.5, and a ratio of negative to positive samples of 2. The calculated sample size was 30, comprising 10 positive (VITT) samples and 20 negative (HIT) samples. Given that VITT is a rare condition with limited sample availability and HIT and healthy control samples are more readily available, for HIT assay, we used 40 samples (20 positive and 20 negative) to enhance statistical power. Serum samples from VITT patients, HIT patients, and healthy controls were collected retrospectively. HIT serum samples were from HIT-suspected patients who tested positive through Lifecodes PF4 IgG HIT ELISA and platelet-based functional assays (Serotonin Release Assay or the PF4-dependent P-Selectin Expression Assay) in the Mayo Clinic Special Coagulation Laboratory. VITT serum samples were collected from VITT-confirmed cases based on the clinical history and laboratory confirmation (Lifecodes PF4 IgG HIT ELISA and PF4-dependent P-Selectin Expression Assay). Control samples were obtained from healthy individuals. Blood samples were obtained with consent from the patient suspected of MGTS and used in research testing. Studies were approved by the Institutional Review Board of Mayo Clinic.

### Materials

4-(2-hydroxyethyl)piperazine-1-ethanesulfonic acid (HEPES), Pb(NO3)2, Ammonium acetate, Tween 20, chlorotrimethylsilane, Amicon Ultra-0.5 Centrifugal Filter, bovine serum albumin, and 6-Mercapto-1-hexanol, sodium chloride (NaCl), 1-ethyl-3-(3-dimethylaminopropyl) carbodiimide (EDC), cysteamine, and dithiothreitol (DTT) were purchased from Sigma-Aldrich (St. Louis, MO, USA). Hydroxy-EG6-undecanethiol was obtained from Dojindo Molecular Technology (Washington, DC, USA). Sodium heparin (Mw: 12 kDa) was purchased from Smithfield Bioscience (Cincinnati, OH, USA). 4-Arm poly (ethylene glycol) thiol (PEG4SH; Mw: 10 kDa) was obtained from Sunbio Inc. (Anyang, Korea). 1 - Hydroxybenzotriazole hydrate (HOBt) was purchased from Anaspec (Fremont, CA, USA). A dialysis membrane was obtained from Spectrum Laboratories (3.5 kDa Mw cut-off, Rancho Dominguez, CA, USA). NHS-activated AuNPs (20 nm) were purchased from CytoDiagnostics (Tulsa, OK, USA). Ethyl alcohol (EtOH) was purchased from Electron Microscopy Sciences (Hat eld, PA, USA) and isopropyl alcohol (IPA) was acquired from Honeywell (Charlotte, NC, USA). Platelet-factor 4 (Human CXCL4) was purchased from Protein Foundry LLC (Milwaukee, WI, USA). Human IgG Antibody was purchased from R&D Systems (Minneapolis, MN, USA). SuperBlock™ Blocking Buffer and phosphate-buffered saline (PBS) were obtained from Thermo Fischer Scientific (Waltham, MA, USA). Silicon wafers were purchased from University Wafer (Boston, MA, USA). SU-8 2025 photoresists, and SU-8 developer were acquired from Kayaku Advanced Materials (Westborough, MA, USA). AZ 5214-E IR photoresist, AZ 300 MIF developer, AZ 10XT photoresist, and AZ-400K developer were purchased from Integrated Micro Materials (Argyle, TX, USA). Poly(dimethylsiloxane) (PDMS) base and curing agent kit (Sylgard 184) were obtained from Ellsworth Adhesives (Minneapolis, MN, USA). Gold etch-type TFA was purchased from Transene Electronic Chemicals (Danvers, MA, USA). CR-7s Chrome Etch was purchased from KMG Electronic Chemicals (Pueblo, CO, USA). Ag/AgCl ink was acquired from CH Instruments (Bee Cave, TX, USA). 3-(3-dimethylaminopropyl) Carbodiimide (EDC), 4-Morpholineethanesulfonic acid (MES), and TRIS buffer were purchased from Millepore Sigma (Burlington, MA, USA).

### Fabrication of electrode arrays

Glass slides were used as substrates for electrode array fabrication. A 10 nm Cr adhesion layer and a 100 nm Au layer were sputter coated onto the glass slides (Lance Goddard Associates, Santa Clara, CA). Next, a 1 μm layer of AZ 5214-E IR photoresist was spin-coated onto the substrate at 5000 rpm and soft baked at 110 °C for 1 min. The desired electrode structures were UV exposure using a micropattern generator (μPG 101, Heidelberg Instruments, Germany) and the exposed photoresist was removed using 300 MIF developer. Subsequently, the patterned Au/Cr layers were etched to create electrodes. To remove any remaining unexposed photoresist, the electrode-patterned slide was sonicated in acetone and then with IPA. Additionally, oxygen plasma treatment was applied for 2 minutes to ensure surface cleanliness. To construct the on-chip Ag/AgCl reference electrodes, Ag/AgCl ink was applied onto the Au electrode surface and cured at 120 °C for 20 minutes.

### Fabrication of molds

The microfluidic devices were designed using CAD software and fabricated using photolithography and soft lithography techniques. Initially, three master molds were fabricated: one for the simple device and two for the automated microfluidic device (a control and a flow layer mold). The simple device mold and the control layer mold of the automated device were fabricated using the same protocol. Firstly, two 4” silicon wafers were spin-coated with negative photoresist (SU8 2025) at 1300 rpm to achieve a 55 μm layer thickness, followed by soft baking on a hot plate at 65 °C for 3 min and 95 °C for 9 min. Subsequently, the photoresists were exposed to UV laser using a micropattern generator (μPG-101, Heidelberg Instruments Inc.). The molds were then baked on a hot plate at 65 °C for 2 min and 95 °C for 7 min, developed by immersion in SU8 developer, and hard baking at 150 °C for 30 min.

To fabricate the flow layer of the automated microfluidic device, a silicon wafer was first coated with a 30 µm thick layer. First, the wafer was spin-coated with two consecutive layers of positive AZ10XT photoresist at 800 rpm with soft bakes of 80 s at 110 °C and 180 s at 115 °C. The structures were then UV exposed and developed with AZ 400k 1:3 developer. A reflow process was performed by baking the mold at 120 °C for 30 min to obtain rounded structures. Then, the temperature was increased to 200 °C with a ramp of 3 °C/min and hard baked for 120 min. Subsequently, the mold was coated with a negative SU8 2025 photoresist layer using the same protocol for the control layer. Finally, all three molds were exposed to chlorotrimethylsilane for one hour.

### Fabrication of PDMS devices

The PDMS base and curing agent were mixed to fabricate the simple devices at a 10:1 ratio (w/w). This mixture was poured into the mold, degassed in a vacuum desiccator for 10 minutes, and baked in a convection oven for 1 hour at 80°C. The cross-linked PDMS was then peeled off from the mold, and the inlets and outlets were punched using a 0.4 μm puncher. Finally, the PDMS devices were treated with oxygen plasma for 2 minutes and aligned onto the array of electrodes.

Automated microfluidic device replicas were fabricated by pouring the PDMS base and curing agent at a 5:1 (w/w) ratio onto the control layer mold and placing them in a vacuum desiccator for 10 minutes. The flow layer mold was spin-coated at 600 rpm with a PDMS-curing agent mix at a 20:1 ratio. Subsequently, both molds were partially cured at 80°C for 20 minutes. Afterward, the control layer replicas were peeled off, cut, and aligned onto the flow layer mold. Both layers were baked together for 90 minutes at 80°C to bond them. Finally, the multilayer devices were peeled off, and the inlets and outlets were punched before bonding the replicas onto the array of electrodes using oxygen plasma treatment.

### Preparation of the AuNPs/anti-human IgG@Pb2+ Immunoprobes

We have previously described a protocol to conjugate AuNPs/Abs@Pb^2+^ (*29*). We followed this protocol for this study to synthesize immunoprobes targeting human IgG (AuNPs/anti-human IgG@Pb2+). Briefly, NHS-activated AuNPs (6.54 × 1011 NPs/mL) were dispersed in 90 μL of 1× PBS containing 40 μg of antibody (Ab) solution and incubated for four hours at room temperature. Thereafter, 10 μL of quencher solution was added to eliminate the remaining unreacted NHS on AuNPs and then incubated with BSA to block the AuNP.

To remove unbound Abs and any reagents, the resulting mixture was centrifuged at 7000g for 20 min and washed three times with ammonium acetate buffer (20mM, pH 6) with 0.05% Tween 20 Next, the synthesized AuNPs/Abs conjugates were dispersed in 0.15 mL of ammonium acetate buffer with 0.025% Tween 20, and incubated with 15 μL of 10 mM Pb(NO3)_2_ of aqueous solution and kept stirring overnight to complex Pb2+ ions with the amine group of Abs. Finally, the AuNPs/Abs@Pb2+ were collected by centrifugation and washed thoroughly with 1 mL of HEPES buffer (0.02 M, pH 7.0) with 0.025% Tween 20.

### Synthesis of heparin-thiol

Thiolated heparin (Hep-SH) was synthesized by converting the carboxyl groups of heparin into thiol groups, as described in a previous report (*42*). Hep-SH with a 30% degree of thiolation was prepared by reacting the carboxyl groups of heparin with cysteamine. Briefly, heparin was dissolved in distilled water (DW) at a concentration of 10 mg/mL. EDC (1.5 molar ratio), HOBT (1.5 molar ratio), and cysteamine (2 molar ratio) were sequentially added, and the pH was adjusted to 6.8. After reaction under stirring for 12 hours at room temperature, the solution was thoroughly dialyzed to remove unreacted reagents. Then, an excess amount of DTT (10 molar ratio) was added to reduce the oxidized disulfide groups and to form free thiol groups on the modified heparin. After 12 hours of reaction, this solution was dialyzed against a 0.1 M NaCl solution at pH 3.5, then at pH 5.0, and finally in pure DIW, sequentially, and then lyophilized. The degree of thiolation was measured using Ellman’s reagent at 412 nm. 30% thiolated heparin was used for all experiments.

### Device setup and operation

First, the microvalves were filled with DI water using Tygon tubing connected to the pneumatic control system delivering a 30-psi pressure. The pneumatic control panel comprised solenoid valves (MH1, Festo) which were controlled with a microcontroller (MEGA 2560, Arduino) via a LabVIEW interface (2023 community edition). The flow layer channels were filled with PBS-T. Afterwards, the microvalves were actuated to isolate the reaction chamber form the central chamber containing the onboard reference/counter electrode. Next, to functionalize the working electrode surfaces, Heparin-SH, PEG-SH, PF4, cross-linked PF4, and PBS-T were loaded into the device and the automated program to surface preparation was activated. To functionalize the working electrode surfaces, the serum samples, anti-human IgG@Pb2+ nanoparticles, and the electrolyte were loaded, and the assay was automatically performed by activating the LabVIEW program. The device uses on-chip peristaltic pumps to draw the reagents and samples into the reaction chambers. The pumps operate at a frequency of 100 ms, achieving a volumetric flow rate of 1 µL/min with a total assay runtime of 50 minutes.

### Functionalization of electrodes and electrochemical detection of HIT and VITT antibodies

The Au working electrodes were functionalized with the antigen targets PF4 or cross-linked PF4 to capture HIT or VITT antibodies. The preparation of the biosensor surface involves initially functionalizing the working electrodes with Heparin-thiol (0.25 mg/ml) by flowing for 10 minutes, followed by washing with phosphate-buffered saline with 0.05% Tween 20 (PBS-T) for 10 minutes. Subsequently, working electrodes are blocked with PEG-thiol (1 mg/ml) by injecting for 10 minutes and then incubating for one hour. After washing, PF4 or cross-linked PF4 (20 μg/ml) was immobilized on the working electrodes. The functionalized electrodes are exposed to serum samples for 10 minutes to detect HIT and VITT antibodies, followed by washing with PBS-T. Next, the anti-human IgG@Pb2+ nanoparticles were injected at a concentration of 6.54E^+11^ particles/mL for 10 minutes. After washing the electrodes with PBS-T and DI water, microvalves to isolate the central chamber were opened to establish an electrolyte connection between the working electrode and working, counter, and reference electrodes. Finally, the device was connected to a potentiostat (PalmSens) and analyzed using square wave voltammetry. The SWV curves were recorded in the electrolyte (acetic acid/sodium acetate buffer; 0.2 M, pH 4.5) in the −0.9 to 0.1 V range (versus Ag/AgCl) with 25 mV amplitude and 15 Hz frequency.

### Surface Plasmon Resonance experiments

We used a surface plasmon resonance (SPR) system (Biosensing Instruments) to confirm surface functionalization steps and to optimize the biosensor surfaces. Au SPR chips were prepared by flowing Heparin-SH (0.25 mg/ml), PEG-SH (1 mg/ml), PF4 (20 μg/ml), serum samples (1:50 dilution in super blocking buffer) and anti-human IgG antibodies (1:100 dilution in PBS). For the optimization of non-specific binding of anti-human IgG@Pb2+ gold nanoparticles, Au SPR chips were incubated for one hour with mercaptohexanol (MCH), polyethylene glycol-thiol (PEG-SH), or hydroxy-EG6-undecanethiol. Then, the anti-human IgG@Pb2+ nanoparticles were injected at 20 μl/min. Resonance units (RU) were acquired from the baseline changes before and after the reagent injection.

### Chemical cross-linking of PF4 polypeptides

For PF4 cross-linking, 1 mg/mL EDC and recombinant PF4 were added at an equal ratio (v:v) in 15 mM MES buffer pH 8.0 for six hours at ambient temperature. Cross-linking reactions were stopped by adding 1M Tris pH 7.4 buffer (i.e., a primary amine-containing buffer) at a 1:4 (v:v) ratio of the cross-linked PF4 polypeptide reaction to Tris buffer.

### Statistical analysis

The datasets were obtained and analyzed using PSTrace5 software to calculate the area under the SWV curves and to obtain the total charge. Prism 6 (GraphPad Software, USA) was utilized for data visualization and statistical analysis, applying the Wilcoxon test for group comparisons and ROC analysis to assess diagnostic performance.

## Data Availability

All data produced in the present study are available upon reasonable request to the authors

## SUPPLEMENTARY MATERIALS

Figures S1 to S9 Table 1

Movie S1

## Acknowledgments

We would like to thank Mayo Clinic’s Advanced Diagnostics Laboratory (ADL) for funding this project. We thank Dr. Hien Le, Dr. Anh Tuan Nguyen, and Dr. Yan Liang for helpful discussions.

## Author contributions

Conceptualization: AR, AP, DFCA, AK.

Methodology: DFCA, AK, SL, AMGS, KG, EM, TQN.

Investigation: DFCA, AK, SL Visualization: DFCA, AK Funding acquisition: AR, AP Supervision: AR, AP

Writing – original draft: DFCA, AK, AR

Writing – review & editing: DFCA, AK, SL, AMGS, KG, TQN, AP, AR.

## Data and materials availability

All data are available in the main text or the supplementary materials.

