## Supplementary material for "Automated microfluidic electrochemical biosensor for the detection of immune-mediated thrombotic disorders": Figures S1 to S9

**Optimization of the electrode interface.**

To optimize our electrochemical biosensor and minimize non-specific interactions, we designed an experiment to evaluate the sensor signal under different conditions: (a) when the electrode surface was fully functionalized with Hep-SH, PEG-SH, PF4, and exposed to HIT serum; (b) when the electrode was fully functionalized with Hep-SH, PEG-SH, PF4, and exposed to control serum; (c) when the electrode contained only Hep and PEG-SH without PF4 or serum; and (d) when the electrode surface had only PEG-SH without heparin, PF4, or serum. As shown in Figure S4, the signal from the electrode with only PEG-SH on the surface was comparable to the signal obtained from the fully functionalized electrode exposed to control serum. This result suggested that the background signal may arise from interactions between the anti-human IgG@Pb^2+^ gold nanoparticles and the gold surface.

 To improve the blocking of the Au surface, we explored two approaches: increasing the incubation time and evaluating three blocking agents: mercaptohexanol (MCH), polyethylene glycol-thiol (PEG-SH), and hydroxy-EG6-undecanethiol. MCH was chosen for its hydrophilic properties that help prevent non-specific binding. PEG-SH was used for its long polyethylene glycol chains, which provide both hydrophilicity and steric hindrance, and hydroxy-EG6-undecanethiol because it forms a highly hydrophilic self-assembled monolayer (SAM) on gold surfaces. We used SPR to analyze the non-specific binding of the anti-human IgG@Pb^2+^ gold nanoparticles (immunoprobes) with the gold surface after incubating with the blocking agents for one hour and with our previous method, which involved flowing PEG-SH for 5 min. The SPR response units (RU) of the immunoprobes were significantly lower (19 RU) when PEG-SH was incubated for 1h compared to PEG-SH flowed for 5 min (165.1 RU), one-hour incubation with MCH (126.6 RU) or hydroxy-EG6-undecanethiol (126.6 RU), Figure S5. Based on these results, we evaluated the signal-to-noise ratio of our electrochemical biosensor when PEG-SH was incubated for 1h. As depicted in Figure S5 SWV curves showed a significant decrease in background signal, yielding an improvement of the signal-to-noise ratio (Q_HIT_/Q_CTL_) from 2.8 to 8.8, Figure S6. Overall, prolonged incubation with PEG-SH proved to be the most effective strategy to minimize nonspecific binding in our electrochemical biosensor.

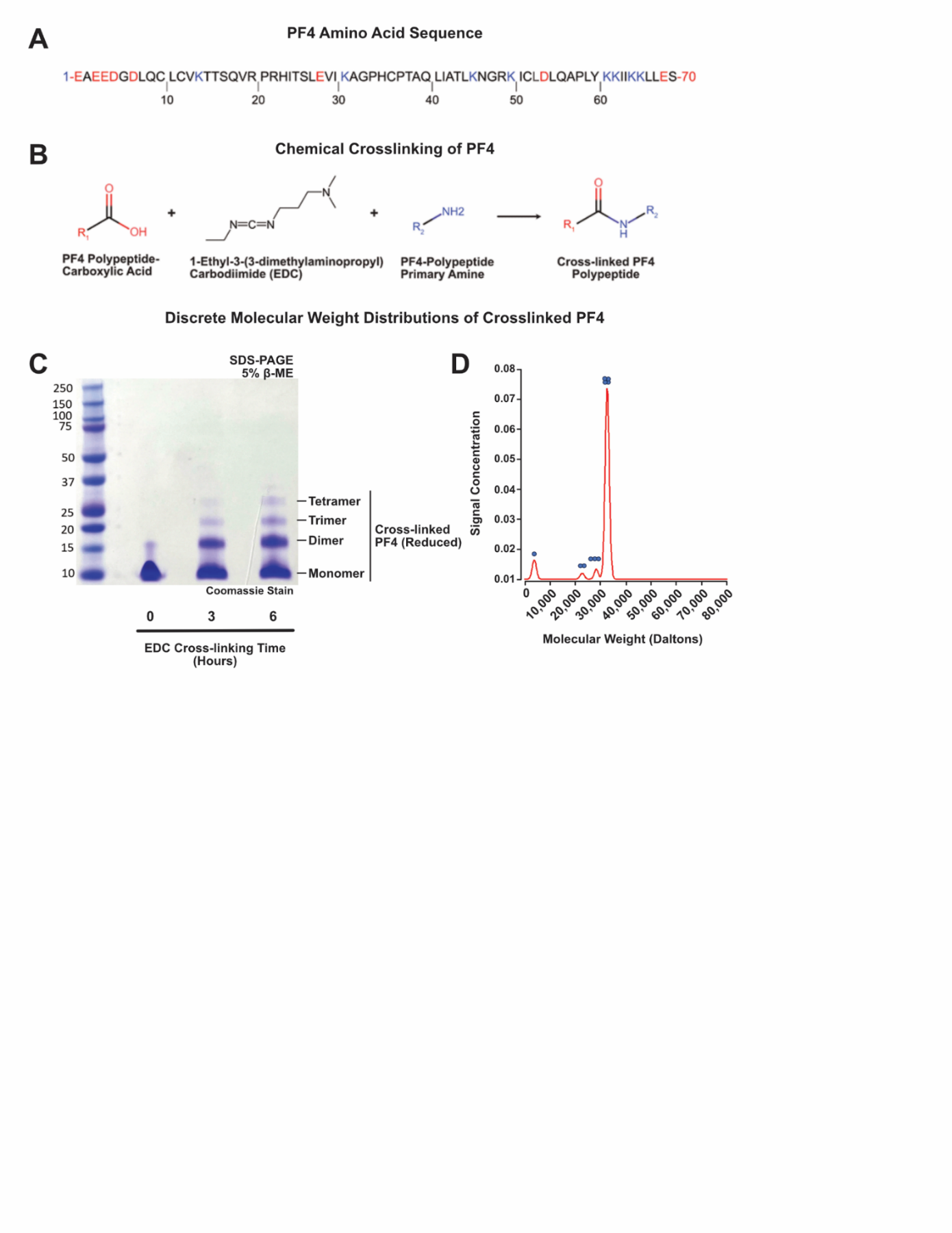

**Figure S1. Recombinant platelet factor 4 was chemically crosslinked as an antigenic target for detecting VITT antibodies.** (**A**) The amino acid sequence of recombinant PF4 polypeptide is shown. Amino acid residues with carboxylic acid groups (and the carboxy terminus of the PF4 polypeptide) are indicated in red. Amino acid residues containing primary amine groups (and the amino terminus of the PF4 polypeptide) are denoted by blue font. (**B**) A schematic diagram for the chemical cross-linking of PF4 polypeptides. (**C**) SDS-PAGE and Coomassie blue staining and (**D**) analytical ultracentrifugation using non-reducing buffer conditions after six hours of EDC-mediated crosslinking.

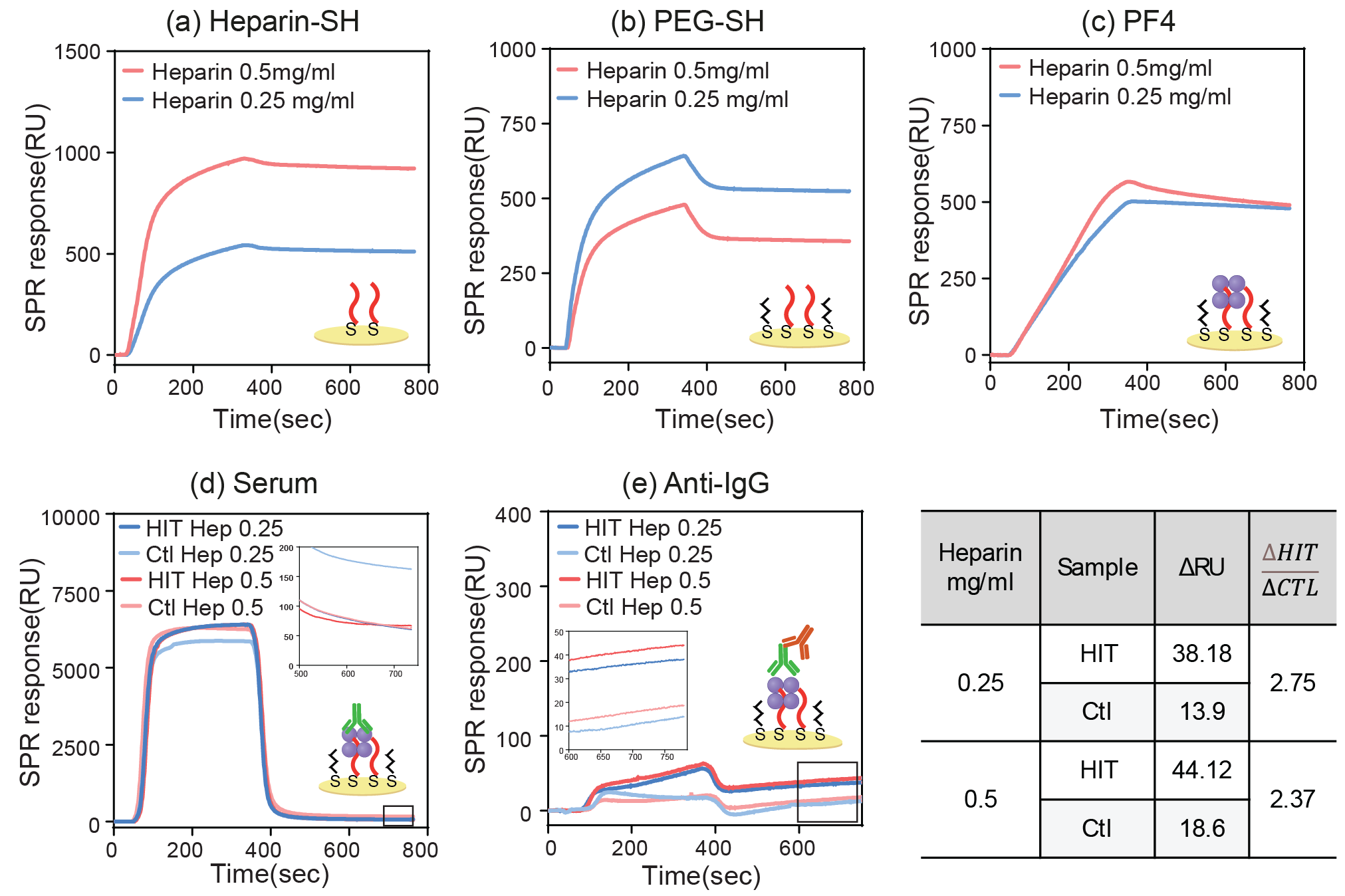
**Figure S2. SPR analysis of biorecognition layer.** (a) Heparin-SH immobilization at two concentrations (0.25 mg/mL and 0.5 mg/mL), (b) PEG-SH immobilization, (c) PF4 binding on heparin-functionalized surfaces, (d) samples from HIT-positive (HIT) and control (Ctl) serum, (e) anti-IgG binding to Heparin/PF4/HIT antibodies complexes. Table summarizes the changes in RU (ΔRU) for HIT and control samples after anti-IgG binding and the signal-to-noise ratio (ΔHIT/ΔCTL).

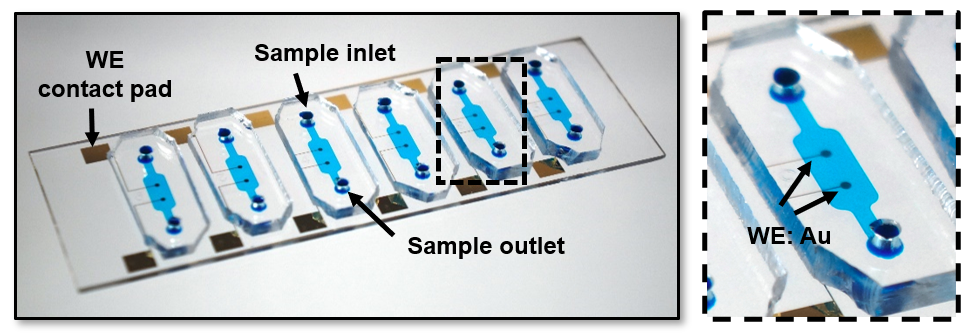

**Figure S3. Photograph of the simplified microfluidic device**. Photograph shows the device with six microchambers filled with blue dye, each chamber featuring an inlet and an outlet for sample introduction, and for connecting off-board reference and counter electrodes. Zoom in on a single microchamber with two working electrodes.

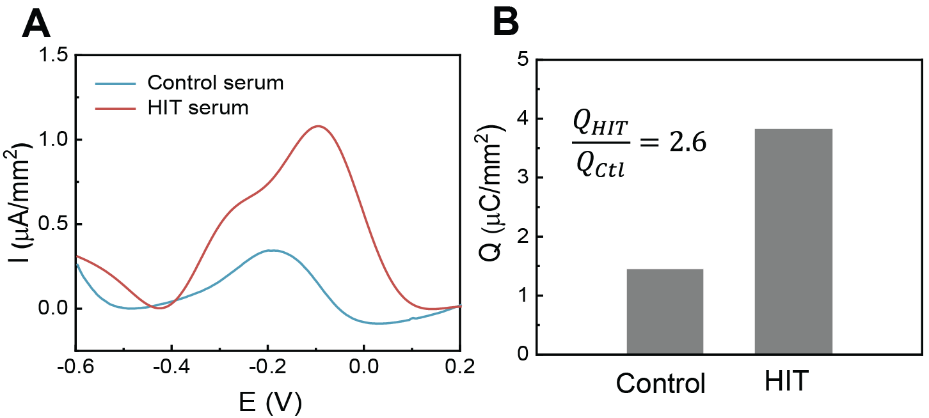

**Figure S4: Electrochemical detection of HIT with a suboptimally passivated biosensor**. (**A**) Square Wave Voltammetry (SWV) signals from healthy donor serum and HIT-positive patient serum, using heparin/PF4 as the antigen target on a suboptimal surface. (**B**) Comparison of the total charge (Q) between healthy donor serum and HIT-positive serum, highlighting the signal-to-noise ratio (Q_HIT_/Q_CTL_).

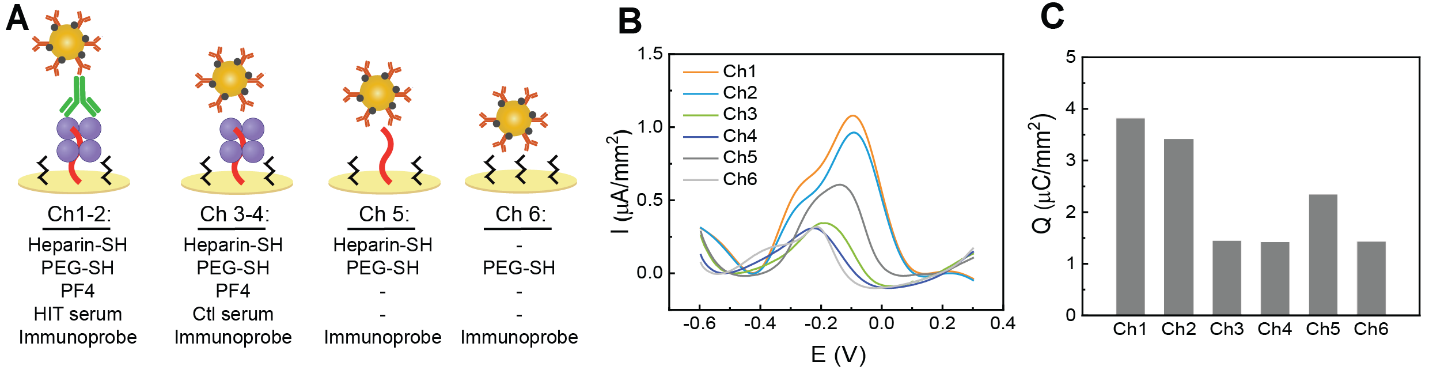

**Figure S5: Characterization of the electrochemical biosensor signal under different conditions**. (**A**) Schematic representation of the surface functionalization for each channel: Ch1-2 are functionalized with Heparin-SH, PEG-SH, and PF4, and exposed to HIT serum and immunoprobe; Ch3-4 are functionalized similarly but exposed to control serum; Ch5 is functionalized with Heparin-SH and PEG-SH but without PF4 or serum; Ch6 is functionalized with PEG-SH only. (**B**) Square Wave Voltammetry (SWV) signals for each channel (Ch1 to Ch6), showing distinct current responses. (**C**) Total charge (Q) calculated for each channel, indicating variability across different surface treatments.

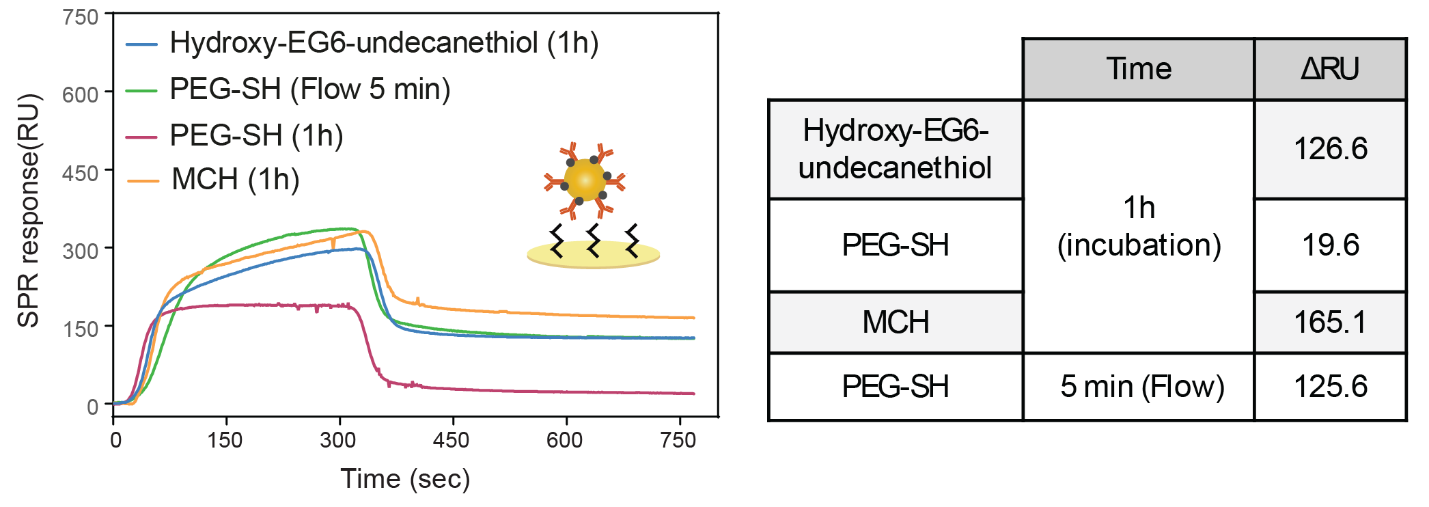
**Figure S6: SPR analysis of gold surface passivation.** Evaluation of the non-specific binding of AuNP/anti-human IgG@Pb^2+^ to the gold surface after treatment with various blocking agents and different flow/incubation times. The table summarizes the changes in response units (ΔRU) for the binding of the immunoprobes.

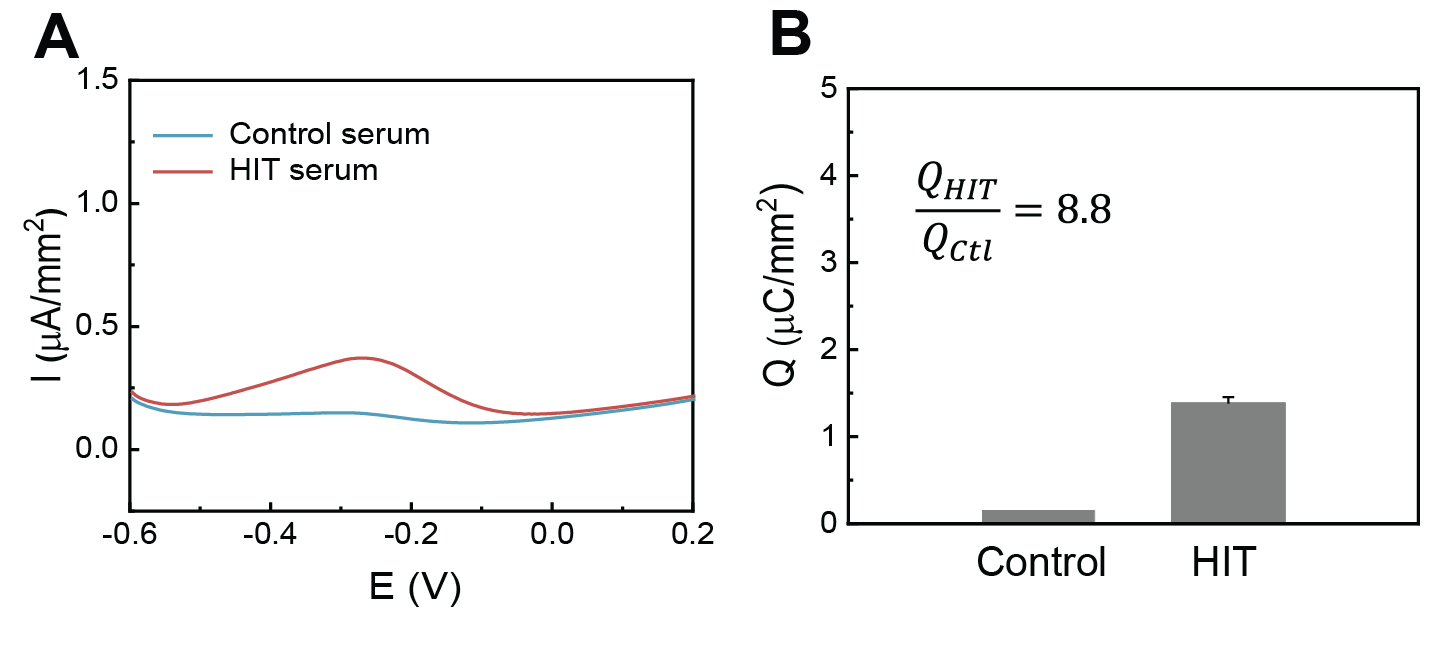

**Figure S7: Electrochemical detection of HIT using an optimal surface**. (**A**) SWV curves from healthy donor serum and HIT serum, using heparin/PF4 as the antigen target on an optimally passivated surface. (**B**) Comparison of the total charge (Q) between healthy donor serum and HIT-positive serum, showing an improved signal-to-noise ratio.

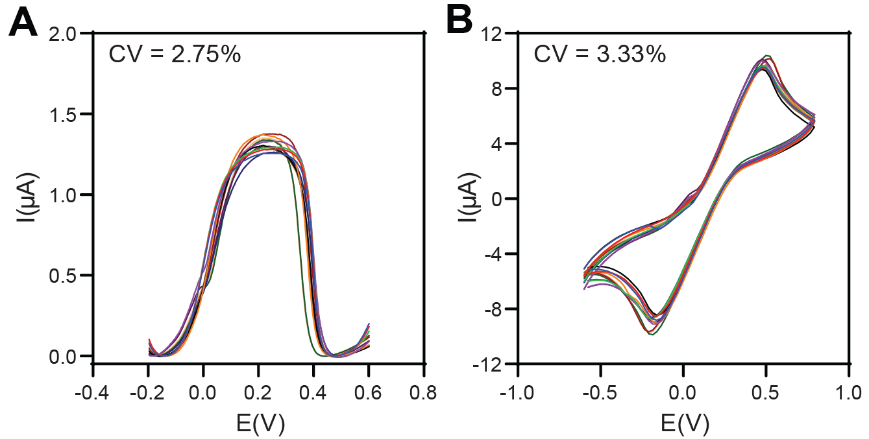

**Figure S8: Electrochemical characterization of the 12 working electrodes**. SWV (**A**) and CV (**B**) curves obtained for the 12 working electrodes connected to the onboard counter and reference electrodes, showing similar signals, measured using 10 mM [Fe(CN)6]4-/3- in 0.1 M KCl.

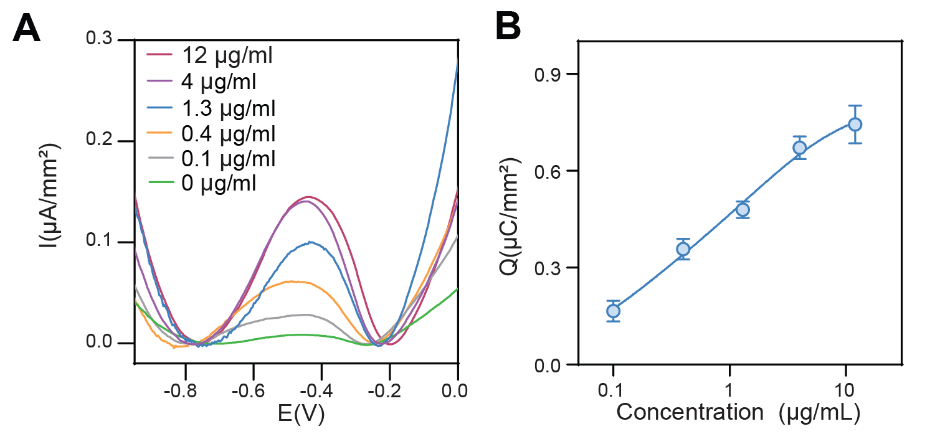

**Figure S9: Evaluation of biosensor responses to HIT-like Abs.** Electrochemical analysis (SWV) of electrodes after incubation with different concentrations of HIT-like antibodies, sequenced and produced from a patient with monoclonal gammopathy of thrombotic significance (MGTS). SWV curves (**A**) and total charge (**B**) show the concentration dependence of the electrochemical signal.
